# Temporal Trends and Demographic Disparities in Percutaneous Left Atrial Appendage Occlusion Utilization in the United States, 2016–2022

**DOI:** 10.64898/2026.10.12.26365359

**Authors:** Brennan Shaun, Mohammad Reza Movahed, Mehrtash Hashemzadeh, Mehrnoosh Hashemzadeh

## Abstract

**Background:** Left atrial appendage occlusion (LAAO) provides non-pharmacological stroke prevention for non-valvular atrial fibrillation patients with contraindications to long-term oral anticoagulation. National long-term data on age-adjusted utilization trends and demographic adoption disparities remain limited.

**Objectives:** To evaluate national temporal trends, hospital characteristics, clinical acuity shifts, and demographic adoption patterns of percutaneous LAAO procedures.

**Methods:** Using the National Inpatient Sample (NIS) from 2016 through 2022, percutaneous LAAO procedures were identified using ICD-10-PCS code 02L73DK. Weighted national estimates were used to evaluate patient demographics, hospital factors, clinical acuity, and overall in-hospital mortality. Age-adjusted rates per 100,000 population were calculated using direct standardization to the 2000 U.S. standard population.

**Results:** A total of 190,020 weighted LAAO procedures were identified increasing from 4,875 in 2016 to 60,445 in 2022. The mean patient age was 76.3 ± 7.6 years, and 58.5% were male. Most patients were White (90.4%), while Hispanic (4.3%), Black (3.9%), and Asian/Pacific Islander (1.3%) patients accounted for smaller proportions of procedures. Most procedures were performed at urban teaching hospitals (87.0%). Overall in-hospital mortality was 0.1% (n = 225, *p* = 0.582). Minor APR-DRG risk of mortality increased from 43.5% to 55.5%, while all others decreased (*p* < 0.001). Age-adjusted LAAO rates rose from 34.19 to 379.87 per 100,000 population (*p*-trend < 0.01). Significant differences in utilization across racial groups persisted throughout the study period.

**Conclusions:** Percutaneous LAAO utilization expanded rapidly with exceptionally low inpatient mortality. Persistent gaps in age-adjusted rates highlight key racial, sex, and geographic disparities in structural heart care.

**Condensed Abstract:** Using the 2016–2022 National Inpatient Sample, this study evaluated temporal trends and demographic adoption patterns of percutaneous left atrial appendage occlusion (LAAO). Among 190,020 weighted procedures, annual volume increased from 4,875 to 60,445. In-hospital mortality was exceptionally low (0.1%). Age-adjusted procedural rates rose from 34.19 to 379.87 per 100,000 population. However, significant disparities persisted, characterized by heavy centralization in urban teaching hospitals (87.0%) and disproportionately lower absolute age-adjusted utilization among female, Black, Hispanic, and Asian/Pacific Islander patients compared to male and White cohorts. Targeted initiatives are needed to ensure equitable access to structural heart interventions.

## Introduction

Atrial fibrillation (AF) is the most common sustained cardiac arrhythmia worldwide and is associated with substantial risk of ischemic stroke, cardiovascular morbidity, and mortality.[1] While oral anticoagulation (OAC) remains the cornerstone of stroke prevention in non-valvular AF, a significant proportion of patients face contraindications, high bleeding risks, or poor tolerance to long-term pharmacotherapy.[2] Left atrial appendage occlusion (LAAO) performed using transcatheter devices such as the Watchman, has emerged as a safe and effective non-pharmacological alternative for stroke risk reduction in these vulnerable populations.[2] Since its initial approval by the U.S. Food and Drug Administration (FDA) in March 2015, the clinical adoption of percutaneous LAAO has expanded across academic and community centers alike.[2,3]

Despite advances in structural heart interventions, differences in the utilization of cardiovascular procedures across demographic groups remain an important healthcare concern in the United States.[4] Historically, racial and ethnic minorities have experienced unequal access to novel structural heart interventions and advanced cardiovascular procedures.[4,5] While single-center studies and early registry analyses have tracked general procedure volumes, comprehensive, nationwide assessments examining long-term demographic adoption patterns and age-adjusted utilization trends following FDA approval remain limited.

Understanding whether procedural adoption varies over time across different demographic and racial groups is critical to identifying potential barriers to access and evaluating public health equity in structural heart care. To address this gap, we used nationally representative inpatient data to examine temporal trends in percutaneous LAAO utilization in the United States from 2016 through 2022, with particular attention to age-adjusted rates across sex, racial, and ethnic groups.

The primary objective of this nationwide study is to evaluate national temporal utilization trends and demographic adoption patterns of percutaneous LAAO procedures. Secondary objectives were to examine patient characteristics, including age, sex, race, primary insurance payer, and income quartile), hospital-level factors (bed size, teaching status, location, and ownership), clinical acuity shifts, and overall in-hospital mortality. We further evaluated whether differences in LAAO utilization across demographic groups changed over time.

## Methods

### Data Source and Study Population

Data were obtained from the Healthcare Cost and Utilization Project (HCUP) National Inpatient Sample (NIS) database spanning calendar years 2016 through 2022. The NIS is the largest publicly available all-payer inpatient healthcare database in the United States, representing a 20% stratified sample of discharge records from U.S. community hospitals. Weighted national estimates were derived using discharge weights provided by HCUP to reflect national procedure estimates. Because the Healthcare Cost and Utilization Project (HCUP) National Inpatient Sample (NIS) is a publicly available, completely de-identified database, this study was deemed not human subjects research and was exempt from formal Institutional Review Board review.

### Identification of LAAO Procedures, Patient Characteristics, and Hospital Characteristics

Annual procedure frequencies for percutaneous LAAO were identified using specific International Classification of Diseases, Tenth Revision, Procedure Coding System (ICD-10-PCS) codes corresponding to transcatheter LAAO (specifically 02L73DK). Demographic variables extracted included age (continuous mean and standard deviation, median, and interquartile range), sex (male and female), race/ethnicity (White, Black, Hispanic, Asian/Pacific Islander), primary expected insurance payer (Medicare, Private, Medicaid, Other), and median household income national quartile based on ZIP code.

Hospital-level characteristics included hospital location and teaching status (urban teaching, urban non-teaching, rural), hospital bed size (small, medium, large), geographic census region (South, Midwest, West, Northeast), and hospital ownership/control (private not-for-profit, investor-owned, government). In-hospital clinical outcomes and patient acuity were evaluated using overall inpatient mortality and the All Patient Refined Diagnosis Related Group (APR-DRG) Risk of Mortality subclass (minor, moderate, major, extreme).

### Statistical Analysis and Standardization

All statistical analyses were conducted using SAS 9.4 (SAS Institute Inc., Cary, NC) and STATA 19 (StataCorp, College Station, TX). Datasets with application of HCUP-provided discharge weights, strata, and cluster variables were applied to account for the complex survey design and generate nationally representative estimates. All reported counts represent weighted estimates of U.S. adult inpatient discharges rather than unique individuals.

Baseline patient demographics, socioeconomic indicators, and hospital-level characteristics were summarized using standard descriptive statistics. Categorical variables were reported as weighted frequencies and percentages, while continuous variables were reported as means with standard deviations (SD) or medians with interquartile ranges (IQR), as appropriate. A complete-case analysis approach was utilized; patients with missing data for primary covariates or outcomes, specifically age, sex, race, or in-hospital mortality, were excluded from the final analytical cohort using listwise deletion.

To evaluate independent post percutaneous left atrial appendage occlusion (LAAO) outcomes, including adult admissions (age ≥50 years) from 2016 to 2022, we constructed a multivariable survey-weighted logistic regression model. The model systematically adjusted for patient age, sex, primary expected payer, median household income quartile of the patient’s ZIP code, and hospital teaching status/location. The independent association of each covariate with the primary outcome was expressed as an adjusted odds ratio (aOR) with a corresponding 95% confidence interval (CI).

To analyze the adjusted temporal trends in mortality across the study period (2016–2022), the admission year was included in the multivariable model as a categorical independent variable. We subsequently employed post-estimation predictive margins to calculate the risk-adjusted predicted probability of mortality for each year. This technique allowed us to estimate the expected mortality rate as if the entire patient cohort were subjected to the specific conditions of a given year, holding all other multivariable covariates constant at their mean distributions. Standard errors and 95% CIs for the predictive margins were calculated utilizing the Delta method.

To account for changes in population structure over time, age-adjusted rates per 100,000 population were computed using the direct standardization method, with the 2000 U.S. standard population as the reference. Age adjustment was performed using standard age groupings consistent with CDC guidelines. Hypothesis testing was two-tailed, and an alpha level of *p* < 0.05 was considered statistically significant for all analyses.

## Results

### Baseline Characteristics of the Study Population

A total of 190,020 patients who underwent Left Atrial Appendage Occlusion (LAAO) between 2016 and 2022 were identified. The mean age of the overall cohort was 76.3 years (SD 7.6), with a median age of 77 years (IQR 72-82). While there was a statistically significant variation in age across the study years (*p* < 0.001), the mean age remained clinically stable, ranging from 75.9 to 76.6 years (Table 1).

**Table 1.**
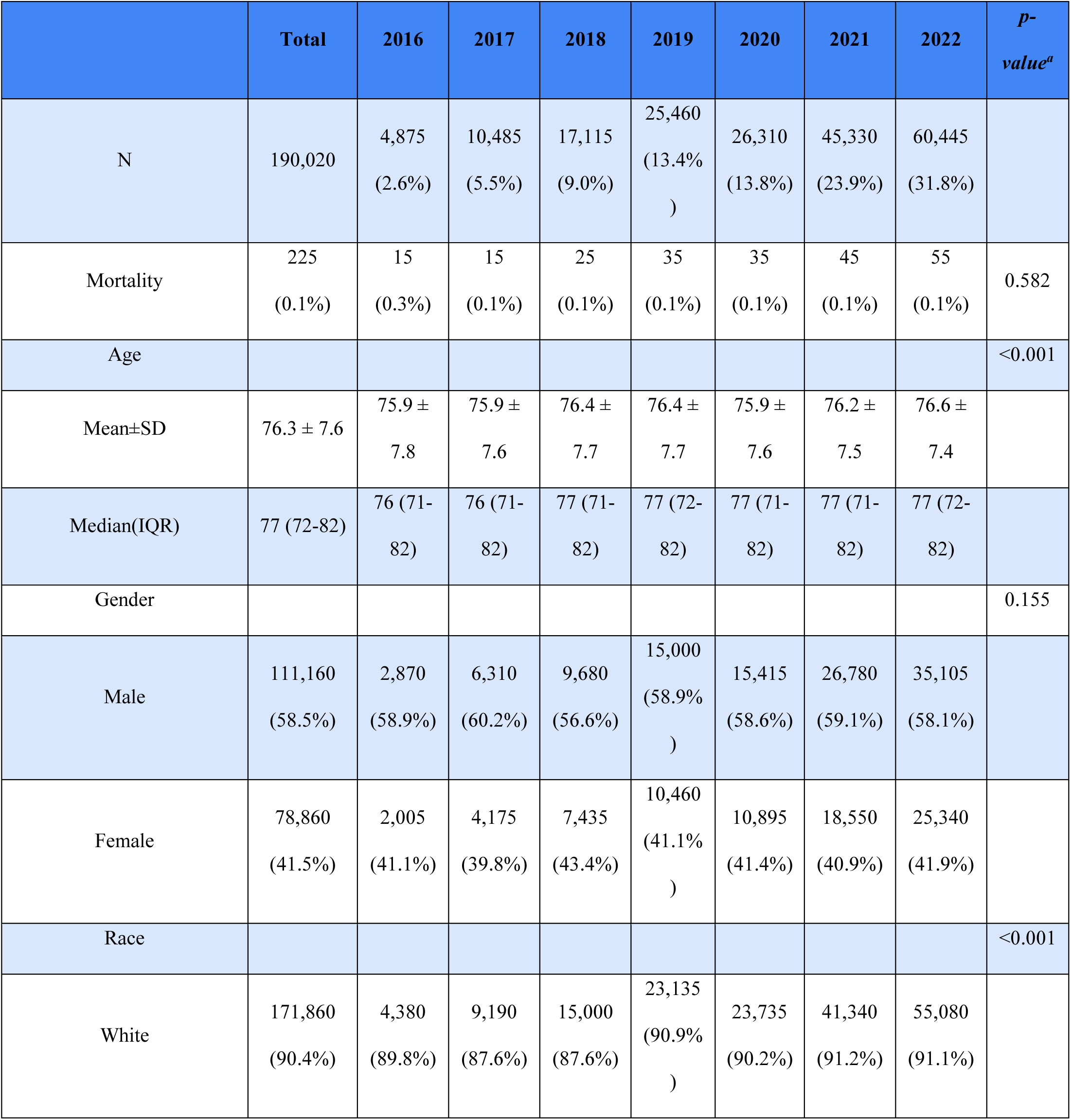
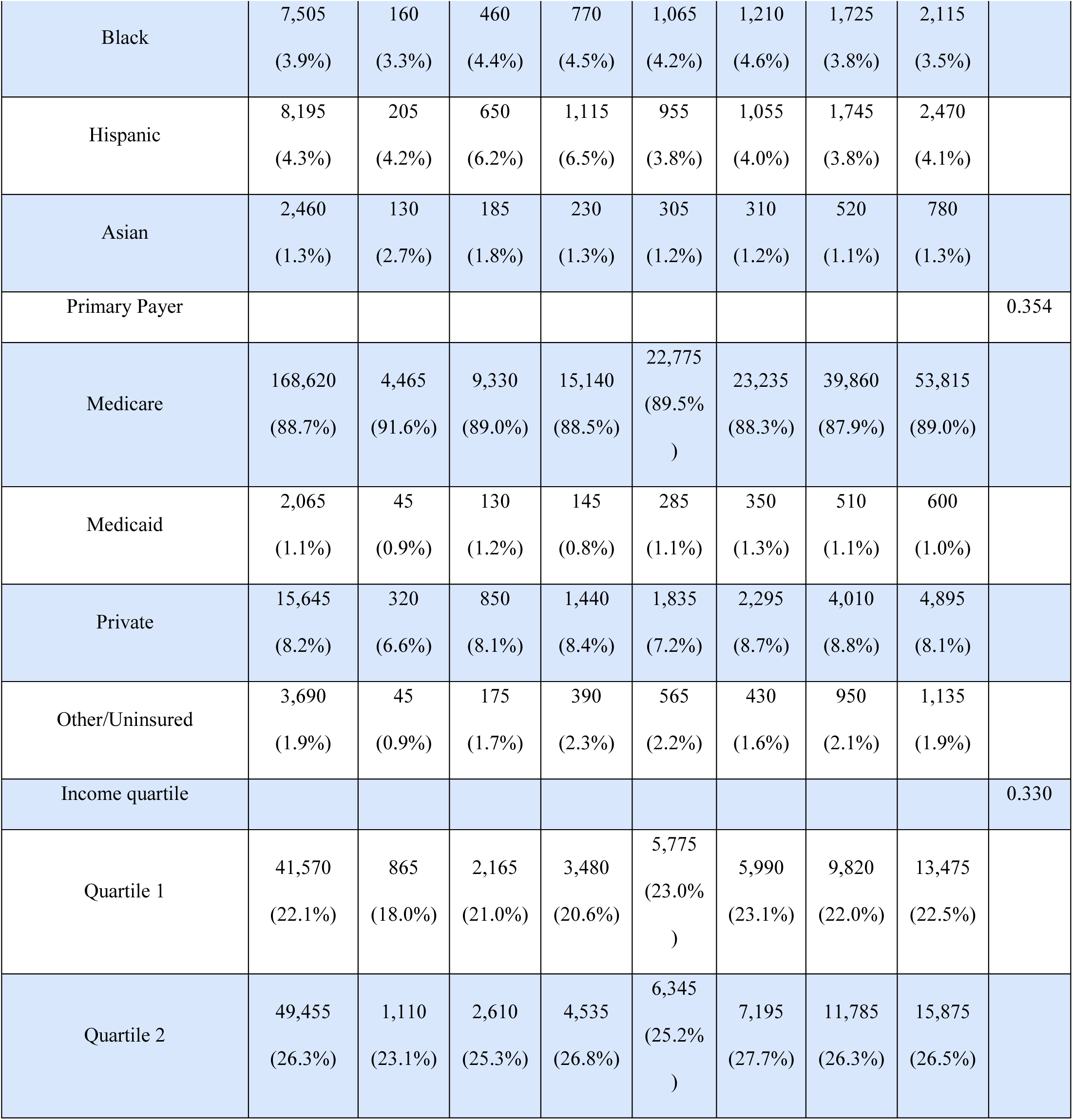
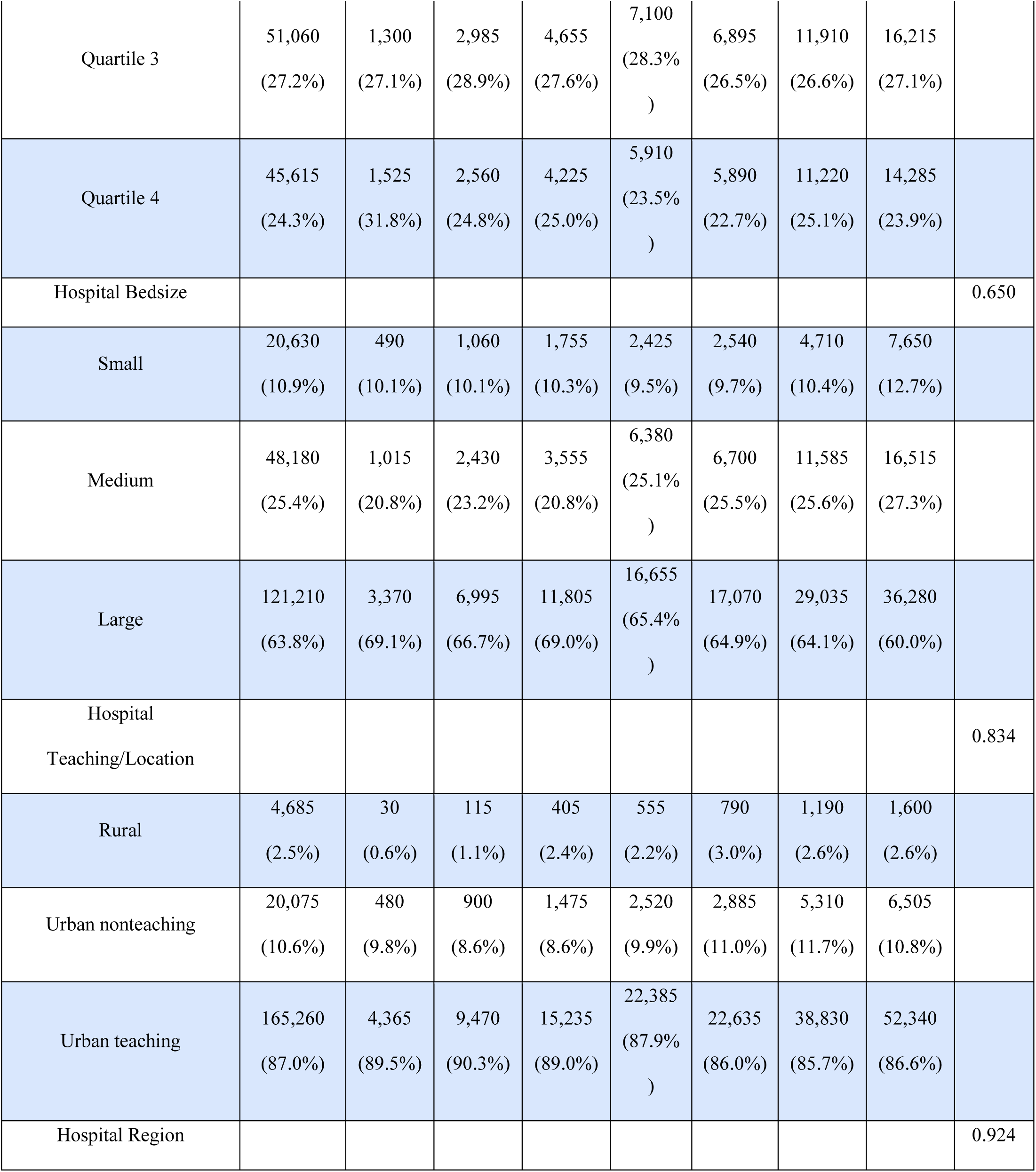
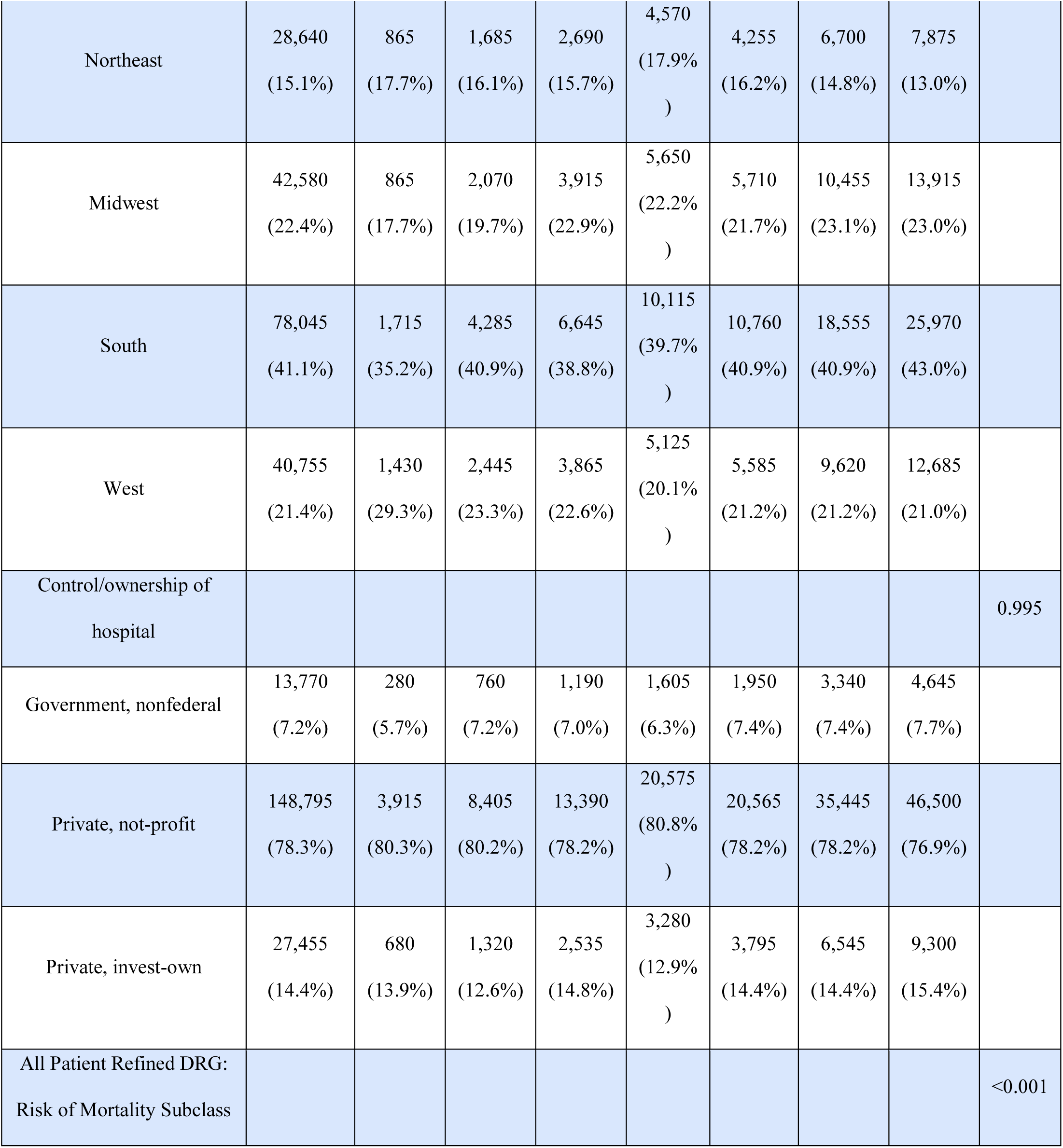
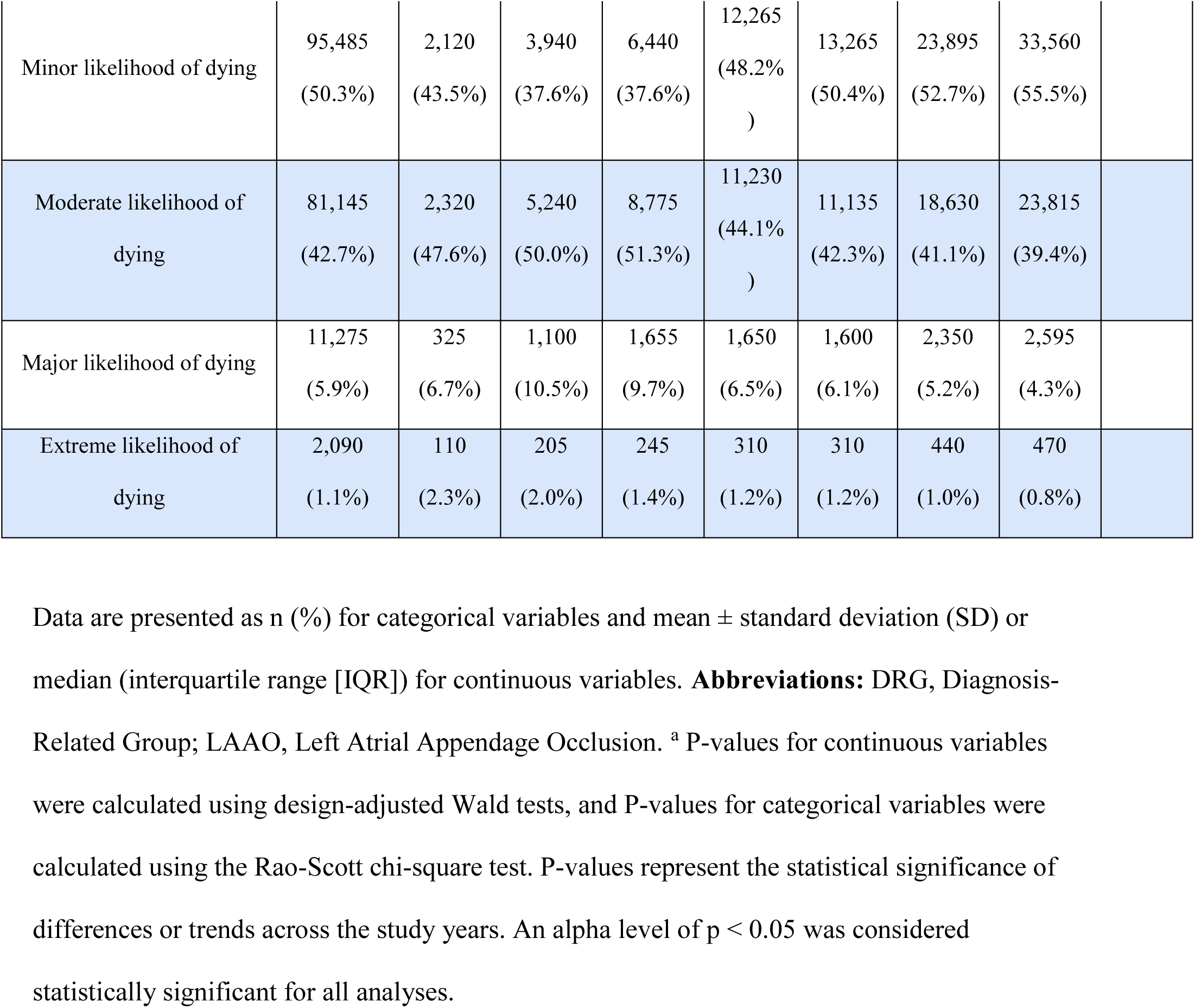
Baseline Patient and Hospital Characteristics of Patients Undergoing LAAO (2016–2022).

The cohort was predominantly male, comprising 58.5% (n=111,160) of the population, compared to 41.5% female (n=78,860). The sex distribution did not change significantly over the study period (*p*=0.155). Analysis of racial demographics demonstrated that the vast majority of LAAO procedures were performed on White patients (90.4%). Minority representation was notably low, consisting of Hispanic (4.3%), Black (3.9%), and Asian/Pacific Islander (1.3%) patients. The racial and ethnic distribution varied significantly across study years (*p* < 0.001) (Table 1).

Socioeconomic and payer data indicated that Medicare served as the primary payer for 88.7% of the cohort, followed by Private insurance (8.2%) and Medicaid (1.1%). Payer distribution remained stable across the study period (*p*=0.354). Patient income was relatively evenly distributed across quartiles, with Quartile 3 representing the largest segment at 27.2%; no significant temporal variation was observed (*p*=0.330) (Table 1).

### Hospital Characteristics

LAAO procedures were predominantly performed at large hospitals, which accounted for 63.8% of procedures, followed by medium-sized hospitals (25.4%) and small hospitals (10.9%). Urban teaching hospitals accounted for 87.0% of all LAAO deployments, whereas rural hospitals accounted for only 2.5%. Geographically, the Southern region performed the most procedures (41.1%), followed by the Midwest (22.4%), West (21.4%), and Northeast (15.1%). Private, not-for-profit institutions represented 78.3% of the hospital control/ownership (Table 1).

### In-Hospital Outcomes

Overall, in-hospital mortality was 0.1% (n=225) and did not change significantly over the study period (*p*=0.582). Patient clinical acuity improved over the study period. According to the All Patient Refined Diagnosis Related Group (APR-DRG) Risk of Mortality subclass, the proportion of patients categorized as having a minor likelihood of dying increased from 43.5% in 2016 to 55.5% in 2022, whereas those classified as having major or extreme mortality risk declined (both *p*<0.001) (Table 1).

### Trends in LAAO Utilization

The weighted national volume of LAAO procedures increased substantially during the study period, from 4,875 procedures in 2016 to 60,445 procedures in 2022. Procedures performed in 2016 accounted for 2.6% of the total weighted volume during the study period, compared with 31.8% in 2022 (Table 2).

**Table 2.**
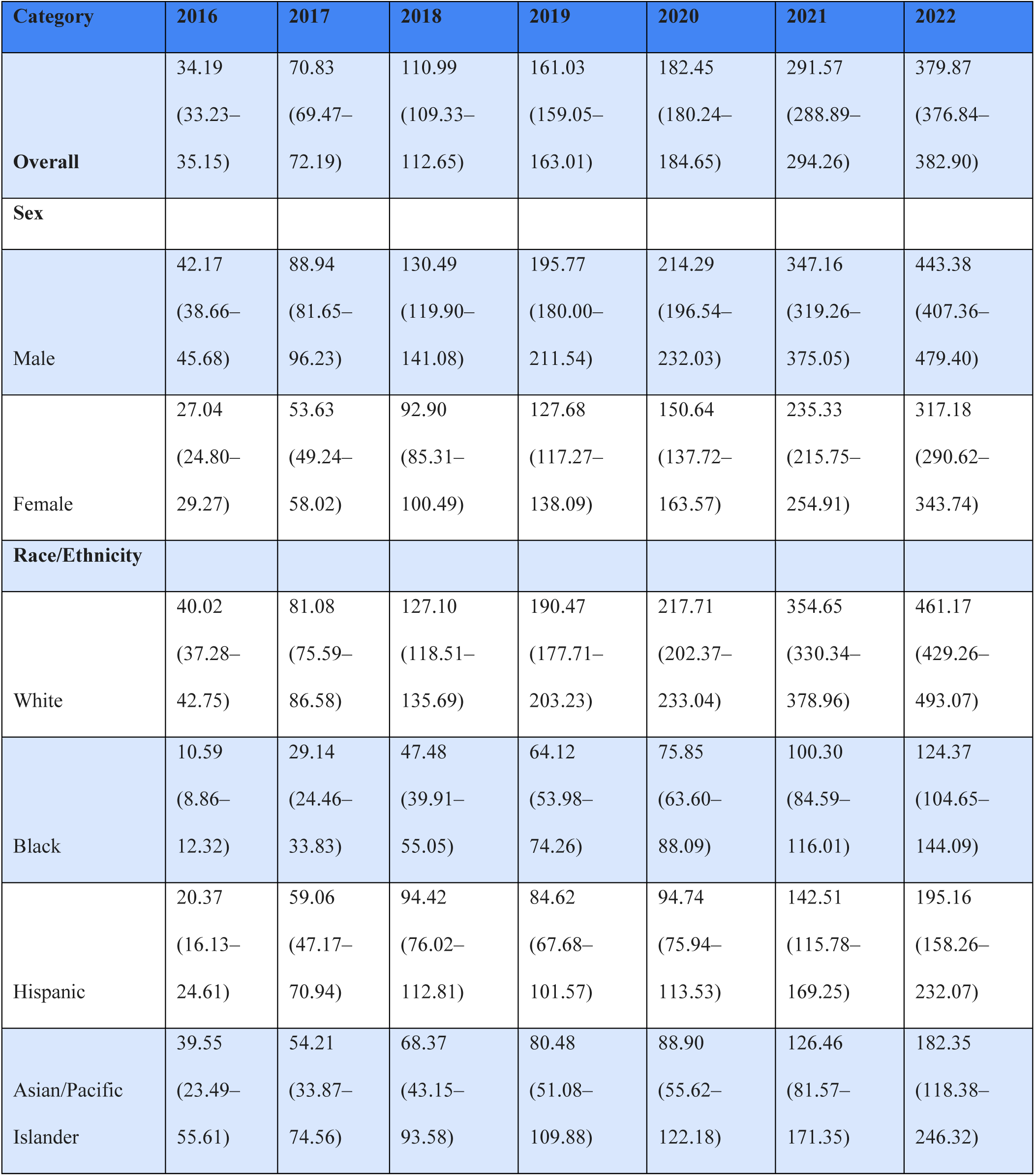
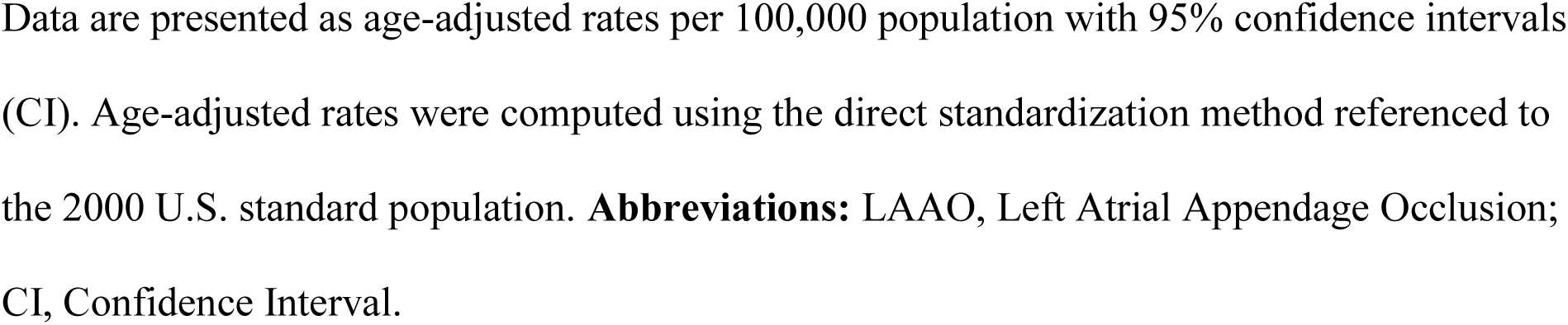
Age-Adjusted Rates of Left Atrial Appendage Occlusion by Sex and Race/Ethnicity (2016–2022).

| Category | 2016 | 2017 | 2018 | 2019 | 2020 | 2021 | 2022 |
| --- | --- | --- | --- | --- | --- | --- | --- |
| **Overall** | 34.19<br>(33.23–<br>35.15) | 70.83<br>(69.47–<br>72.19) | 110.99<br>(109.33–<br>112.65) | 161.03<br>(159.05–<br>163.01) | 182.45<br>(180.24–<br>184.65) | 291.57<br>(288.89–<br>294.26) | 379.87<br>(376.84–<br>382.90) |
| **Sex** |  |  |  |  |  |  |  |
| Male | 42.17<br>(38.66–<br>45.68) | 88.94<br>(81.65–<br>96.23) | 130.49<br>(119.90–<br>141.08) | 195.77<br>(180.00–<br>211.54) | 214.29<br>(196.54–<br>232.03) | 347.16<br>(319.26–<br>375.05) | 443.38<br>(407.36–<br>479.40) |
| Female | 27.04<br>(24.80–<br>29.27) | 53.63<br>(49.24–<br>58.02) | 92.90<br>(85.31–<br>100.49) | 127.68<br>(117.27–<br>138.09) | 150.64<br>(137.72–<br>163.57) | 235.33<br>(215.75–<br>254.91) | 317.18<br>(290.62–<br>343.74) |
| **Race/Ethnicity** |  |  |  |  |  |  |  |
| White | 40.02<br>(37.28–<br>42.75) | 81.08<br>(75.59–<br>86.58) | 127.10<br>(118.51–<br>135.69) | 190.47<br>(177.71–<br>203.23) | 217.71<br>(202.37–<br>233.04) | 354.65<br>(330.34–<br>378.96) | 461.17<br>(429.26–<br>493.07) |
| Black | 10.59<br>(8.86–<br>12.32) | 29.14<br>(24.46–<br>33.83) | 47.48<br>(39.91–<br>55.05) | 64.12<br>(53.98–<br>74.26) | 75.85<br>(63.60–<br>88.09) | 100.30<br>(84.59–<br>116.01) | 124.37<br>(104.65–<br>144.09) |
| Hispanic | 20.37<br>(16.13–<br>24.61) | 59.06<br>(47.17–<br>70.94) | 94.42<br>(76.02–<br>112.81) | 84.62<br>(67.68–<br>101.57) | 94.74<br>(75.94–<br>113.53) | 142.51<br>(115.78–<br>169.25) | 195.16<br>(158.26–<br>232.07) |
| Asian/Pacific<br>Islander | 39.55<br>(23.49–<br>55.61) | 54.21<br>(33.87–<br>74.56) | 68.37<br>(43.15–<br>93.58) | 80.48<br>(51.08–<br>109.88) | 88.90<br>(55.62–<br>122.18) | 126.46<br>(81.57–<br>171.35) | 182.35<br>(118.38–<br>246.32) |

Data are presented as age-adjusted rates per 100,000 population with 95% confidence intervals (CI). Age-adjusted rates were computed using the direct standardization method referenced to the 2000 U.S. standard population. **Abbreviations:** LAAO, Left Atrial Appendage Occlusion; CI, Confidence Interval.

### Age-Adjusted Trends in Left Atrial Appendage Occlusion

The age-adjusted national rate of left atrial appendage occlusion (LAAO) increased steadily throughout the study period from 34.19 per 100,000 population (95% CI, 33.23–35.15) in 2016 to 379.87 per 100,000 population (95% CI, 376.84–382.90) in 2022, representing an approximately 11-fold increase over seven years. Annual age-adjusted rates increased consistently, reaching 70.83 (95% CI, 69.47–72.19) in 2017, 110.99 (95% CI, 109.33–112.65) in 2018, 161.03 (95% CI, 159.05–163.01) in 2019, 182.45 (95% CI, 180.24–184.65) in 2020, and 291.57 (95% CI, 288.89–294.26) per 100,000 population in 2021 before reaching the highest observed rate in 2022. The largest absolute annual increase occurred between 2020 and 2021, when the age-adjusted rate increased by 109.12 procedures per 100,000 population. The increasing age-adjusted utilization paralleled the rise in weighted national procedure volume, which increased from 4,875 procedures in 2016 to 60,445 procedures in 2022, despite relatively stable annual study populations (Table 2, Figure 1).

**Figure 1.**
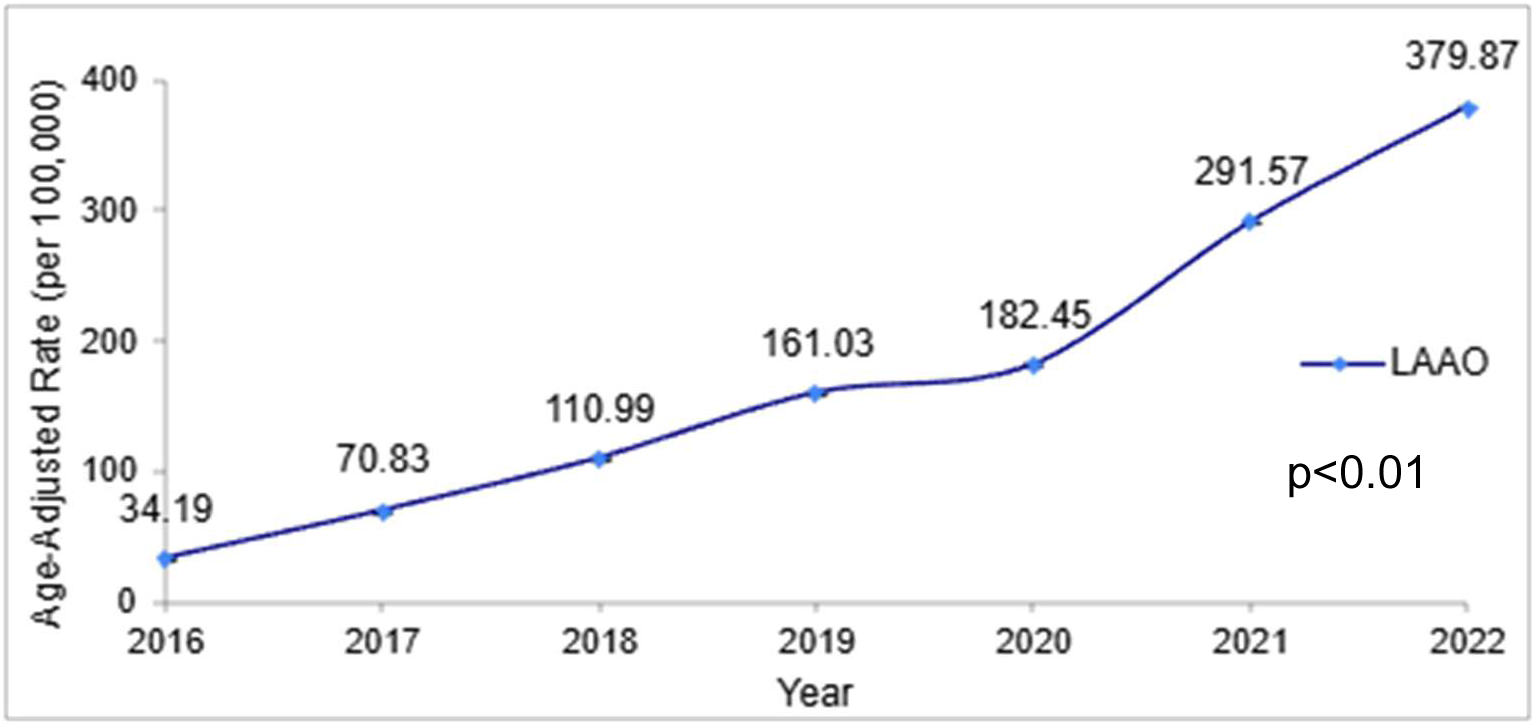
Temporal Trends in Age-Adjusted Rates of Left Atrial Appendage Occlusion (2016–2022). Annual age-adjusted rates of LAAO per 100,000 population. Age-adjusted rates were computed using the direct standardization method referenced to the 2000 U.S. standard population. The P-value (P < 0.01) indicates a statistically significant temporal trend over the study period. **Abbreviations:** LAAO, Left Atrial Appendage Occlusion.

### By Sex

Between 2016 and 2022, national age-adjusted LAAO rates per 100,000 population increased significantly across both sex cohorts. Male patients maintained higher overall procedural rates throughout the study period, rising from 42.17 in 2016 to 443.38 in 2022 (+401.21; 10.5-fold increase). Among female patients, age-adjusted rates expanded from 27.04 in 2016 to 317.18 in 2022 (+290.14; 11.7-fold increase). Across all analyzed study years, male patients consistently demonstrated higher absolute procedural volumes and higher age-adjusted rates compared to female patients (Table 2, Figure 2).

**Figure 2.**
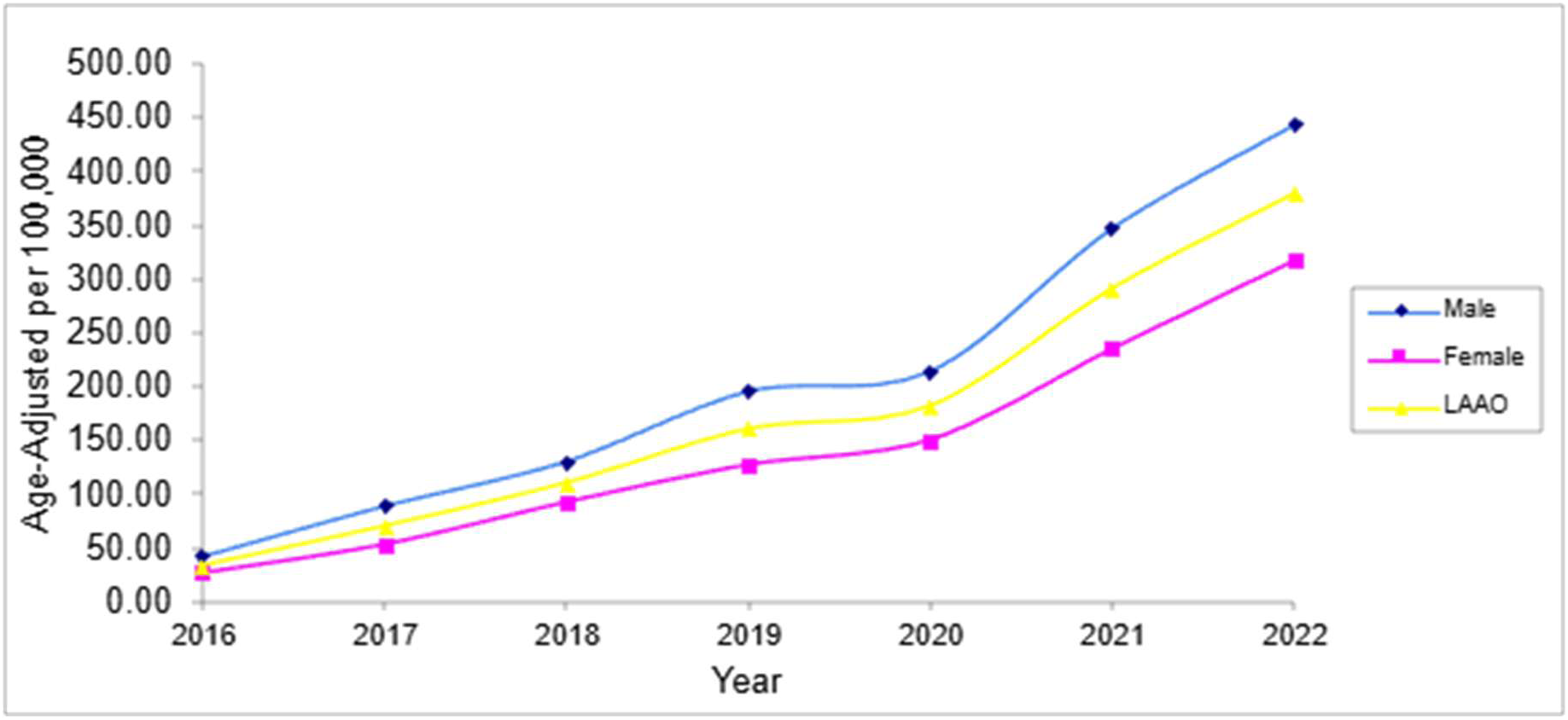
Sex-Based Differences in Temporal Trends of Age-Adjusted Left Atrial Appendage Occlusion Rates (2016–2022). Annual age-adjusted rates of LAAO per 100,000 population, stratified by sex. Age-adjusted rates were computed using the direct standardization method referenced to the 2000 U.S. standard population. The blue line represents male patients, the pink line represents female patients, and the yellow line represents the overall LAAO cohort. Abbreviations: LAAO, Left Atrial Appendage Occlusion.

### By Race

Between 2016 and 2022, national age-adjusted LAAO rates per 100,000 population increased across all racial groups. White patients maintained the highest overall rate, rising from 40.02 in 2016 to 461.17 in 2022 (+421.15; 11.5-fold increase). Among minority cohorts, Hispanic patients experienced the largest absolute increase, rising from 20.37 to 195.16 (+174.79; 9.6-fold increase). Asian/Pacific Islander patients increased from 39.55 to 182.35 (+142.80; 4.6-fold increase), while Black patients expanded from 10.59 to 124.37 (+113.78; 11.7-fold increase) (Table 2, Figure 3).

**Figure 3.**
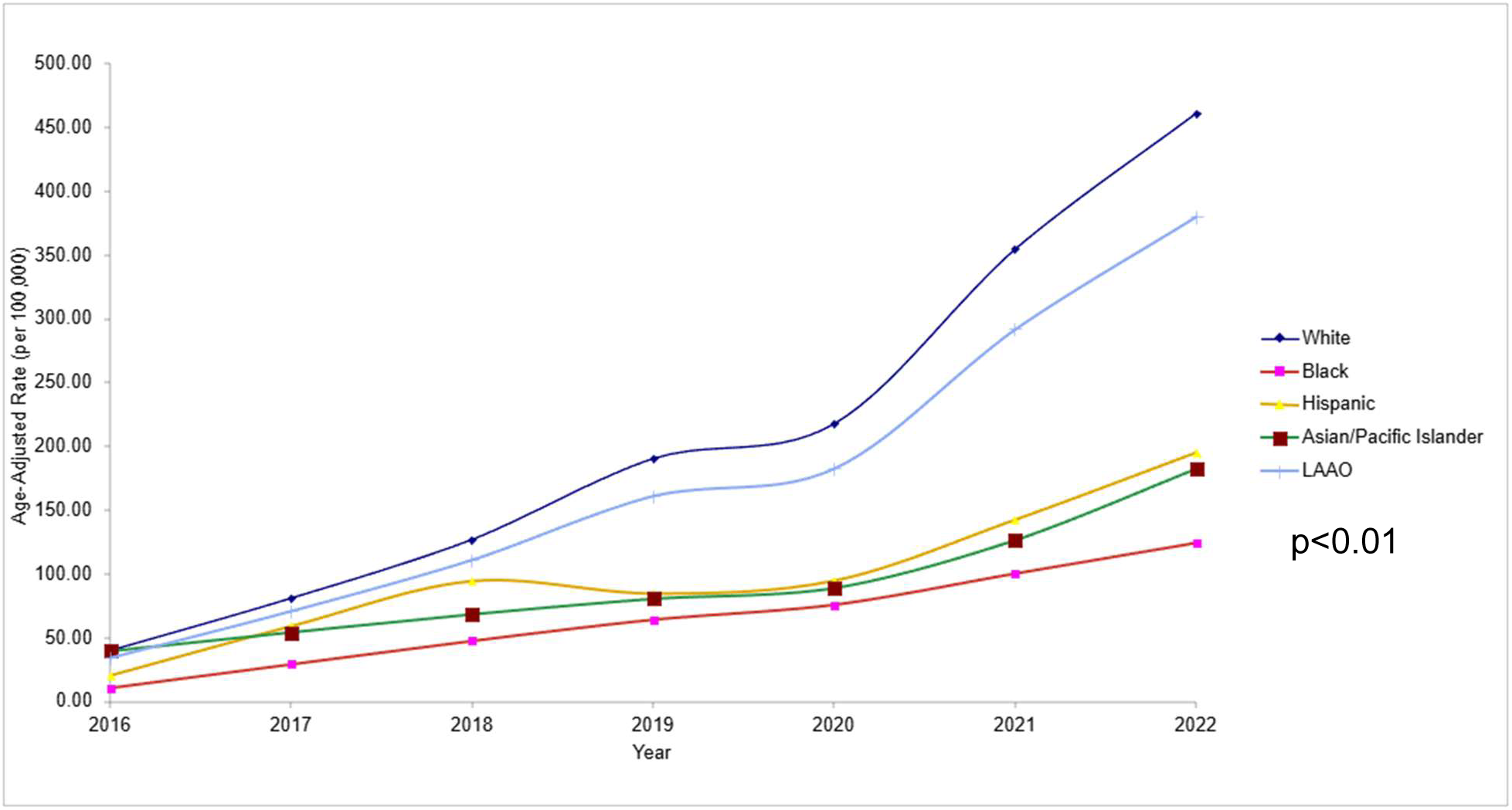
Racial and Ethnic Differences in Temporal Trends of Age-Adjusted Left Atrial Appendage Occlusion Rates (2016–2022). (Central Illustration) Annual age-adjusted rates of LAAO per 100,000 population, stratified by race and ethnicity. Age-adjusted rates were computed using the direct standardization method referenced to the 2000 U.S. standard population. The P-value (P < 0.01) indicates a statistically significant temporal trend over the study period. **Abbreviations:** LAAO, Left Atrial Appendage Occlusion.

## Discussion

This nationwide study provides a comprehensive analysis of baseline clinical characteristics, hospital structural environments, in-hospital outcomes, and age-adjusted temporal utilization trends for percutaneous LAAO in the United States from 2016 to 2022. Our findings demonstrate a rapid expansion in annual procedural volume following FDA clearance for the Watchman device, reaching 60,445 annual events by 2022, with the Southern region consistently performing the highest proportion of procedures throughout the study period. However, the study highlights persistent demographic disparities, characterized by disproportionately higher age-adjusted adoption rates among White and male patients, as well as a heavy centralization of procedures within large urban teaching institutions.

The structural profile reveals no statistically significant shift in hospital location, bed size, or census region between 2016-2022. In total, 87.0% of procedures occurred in urban teaching facilities, with rural settings accounting for only 2.5% of cases. This difference highlights consistent foundational access inequities that prevent more widespread clinical adoption.

Regarding race, statistically significant changes over time were noted, and while overall procedural rates increased, large relative and absolute disparities persisted. Overall White patients accounted for 90.4% of total procedures, whereas Black (3.9%), Hispanic (4.3%) and Asian/Pacific Islander (1.3%) patients comprised a minor fraction. These marked disparities remained after age-adjustment analysis. These findings also highlight underlying systemic barriers, such as referral networks and structural access to specialized centers performing LAAO procedures, echoing patterns of racial underrepresentation and heavy urban centralization documented in earlier literature in both 2019 and 2023. [4,5]

Sex-specific analyses demonstrated a similar pattern. Female patients experienced a greater relative increase in age-adjusted utilization over the study period than male patients (11.7-fold vs. 10.5-fold for males). However, their absolute age-adjusted procedural rates began lower in 2016 and remained substantially lower than those of male patients in 2022 (317.18 vs 443.38 per 100,000 population). This pattern reflects that while females experienced a higher relative rate of expansion, their initial utilization was lower, and absolute rates continue to lag behind males. The age-adjusted utilization gap between men and women echoes sex-based disparities documented across transcatheter structural heart interventions.[6]

There was a statistically significant difference in age across the years studied; however, it was not clinically significant. The mean age shifted slightly from 75.9 ± 7.8 years in 2016 to 76.6 ± 7.4 years in 2022, with an overall mean of 76.3 ± 7.6 years. Median household income quartiles were distributed relatively evenly across all four national household income quartiles (ranging from 22.1% in the lowest quartile to 27.2% in the third quartile). The lack of a pronounced income-based gradient suggests that access to LAAO is relatively equitable across economic strata. This balanced distribution is largely facilitated by the predominant reliance on Medicare coverage (88.7% of the total cohort), which provides a standardized reimbursement for stroke prevention in nonvalvular AF and mitigates direct income-based barriers for patients who are successfully screened and referred.

Notably, in-hospital mortality remained exceptionally low (0.1%), accompanied by a favorable shift toward lower patient clinical risk profiles (APR-DRG minor risk increasing to 55.5% in 2022). This steady improvement in baseline APR-DRG risk profiles over time suggests growing operator experience, device iterations (such as the FDA-approval of the second-generation Watchman FLX technology in July 2020), and expanding operator comfort in treating patients earlier in their disease course. These national observations align with benchmarks from the National Cardiovascular Data Registry (NCDR) LAAO Registry, which documented progressive procedural safety improvements coinciding with operator maturation and next-generation device deployment. [3,7]

Overall, the marked concentration of LAAO procedures within urban teaching hospitals (87.0%) alongside minimal adoption in rural centers (2.5%) highlights a profound geographic centralization of structural heart interventions. When combined with observed sex and racial disparities, these regional patterns suggest that healthcare access gaps extend beyond financial coverage. Minority cohorts residing in rural or medically underserved areas face multiple barriers including, but not limited to, geographic distance to tertiary EP centers, referral bottlenecks from community providers, and indirect social determinants of health (e.g., transportation and caregiver support). All these findings highlight the importance of addressing these persistent inequities and require targeted strategies to further identify areas/barriers restricting equitable access, and implementing interventions such as expanding regional referral networks, enhancing community-level AF screening to facilitate equal access to specialized structural heart care.

### Limitations

This study has limitations inherent to administrative database analyses. First, our data relies on ICD-10 coding, which may be subject to coding errors or misclassification. Second, because the NIS database captures inpatient admissions, procedures performed in ambulatory or outpatient observation settings, something that has expanded in recent years, may not be fully captured. Thus, potentially underestimating total national volume in later study years.

Finally, as an epidemiological trend study, the NIS does not provide detailed patient-level clinical information needed to evaluate individual clinical decision-making components, detailed anatomical considerations, height, weight, specific catheter sizes, precise medication doses, operator volume, or the long-term outcomes after discharge could not be evaluated. The database also does not allow assessment of referral patterns, treatment eligibility among patients who did not undergo LAAO, patient preferences, or other factors that may contribute to observed differences in utilization. Additionally, because the NIS is a discharge-level database, individual patients cannot be followed longitudinally across hospitalizations, limiting assessment of long-term outcomes and repeat hospitalizations.

### Conclusion

Percutaneous left atrial appendage occlusion utilization increased substantially in the United States between 2016 and 2022, with an estimated 190,020 weighted inpatient LAAO discharges during the study period and low inpatient mortality rate of 0.1%. While procedure adoption expanded universally, persistent gaps in age-adjusted rates highlight significant racial/ethnic, sex, and geographic disparities in structural heart procedure adoption. Targeted equity initiatives are essential to ensure broad access to advanced non-pharmacological stroke prevention therapies for patients with non-valvular atrial fibrillation that are candidates for LAAO.

## Disclosures

Nothing to declare.

## Data Availability

NIS is a public database

## Abbreviations

AF: Atrial fibrillation
aOR: Adjusted odds ratio
APR-DRG: All Patient Refined Diagnosis Related Group
CI: Confidence interval
FDA: Food and Drug Administration
HCUP: Healthcare Cost and Utilization Project
ICD-10-PCS: International Classification of Diseases, Tenth Revision, Procedure Coding System
IQR: Interquartile range
LAAO: Left atrial appendage occlusion
NCDR: National Cardiovascular Data Registry
NIS: National Inpatient Sample
OAC: Oral anticoagulation
SD: Standard deviation

## Notes

### Competing Interest Statement

The authors have declared no competing interest.

### Author Declarations

This study is IRB exempt

